# PET/CT and SPECT/CT imaging of ^90^Y hepatic radioembolization at therapeutic and diagnostic activity levels: anthropomorphic phantom study

**DOI:** 10.1101/2022.07.07.22277361

**Authors:** A Budzyńska, A Kubik, K Kacperski, P Szubstarska, M Kuć, P Piasecki, M Konior, M Gryziński, M Dziuk, E Iller

## Abstract

**Purpose:** Prior to ^90^Y radioembolization procedure a pretherapy simulation using ^99m^Tc-MAA is performed. Alternatively, a small dosage of ^90^Y microspheres could be used. We aimed to assess the accuracy of lung shunt fraction (LSF) estimation in both high activity ^90^Y posttreatment and pretreatment scans with isotope activity of ∼100 MBq, using different imaging techniques. Additionally, we assessed the feasibility of visualising hot and cold hepatic tumours in PET/CT and Bremsstrahlung SPECT/CT images.

**Materials and Methods:** Anthropomorphic phantom including liver (with two spherical tumours) and lung inserts was filled with ^90^Y chloride to simulate an LSF of 9.8%. The total initial activity in the liver was 1451 MBq, including 19.4 MBq in the hot sphere. Nine measurement sessions including PET/CT, SPECT/CT, and planar images were acquired at activities in the whole phantom ranging from 1618 MBq down to 43 MBq.

The visibility of the tumours was appraised based on independent observers’ scores. Quantitatively, contrast-to-noise ratio (CNR) was calculated for both spheres in all images.

**Results:** LSF estimation: For high activity in the phantom, PET reconstructions slightly underestimated the LSF; absolute difference was <1.5pp (percent point). For activity <100 MBq, the LSF was overestimated. Both SPECT and planar scintigraphy overestimated the LSF for all activities.

Foci visibility: For SPECT/CT the cold tumour proved too small to be discernible (CNR <0.5) regardless of the ^90^Y activity in the liver, while hot sphere was visible for activity >200 MBq (CNR>4). For PET/CT, the cold tumour was only visible with the highest ^90^Y activity (CNR>4), whereas the hot one was seen for activity >100 MBq (CNR>5).

**Conclusions:** PET/CT may accurately estimate the LSF in a ^90^Y posttreatment procedure. However, at low activities of about 100 MBq it seems to provide unreliable estimations. PET imaging provided better visualisation of both hot and cold tumours.

## Introduction

Radioembolization is a method of hepatic tumours treatment where microspheres containing Yttrium-90 are administered into the arterial vasculature of the liver, to be delivered into close proximity of the tumour. The tumour is then irradiated by β^−^ particles emitted in ^90^Y decay (1).

Prior to radiation microsphere therapy, patients undergo relevant planning studies, including mapping angiography and ^99m^Tc-labeled macroaggregated albumin (^99m^Tc-MAA) imaging (2). A diagnostic dose of ^99m^Tc-MAA is injected through hepatic arteries, just like ^90^Y microspheres. This pretherapy simulation is performed to predict ^90^Y microsphere distribution (3). One of the main goals of the ^99m^Tc-MAA examination concerns the issue of the safety of the planned therapy: estimation of the lung shunt fraction (LSF) as well as detection of potential extrahepatic depositions (4,5). This is because radiation induced pneumonitis and sclerosis due to hepatopulmonary shunting of ^90^Y microspheres is a major toxicity concern in radioembolization procedure (6).

Currently, in most of the nuclear medicine facilities that provide radioembolization treatment, LSF is routinely estimated by ^99m^Tc-MAA planar scintigraphy performed without accounting for attenuation or scatter effects. Because lung and liver have different tissue densities, the LSF will be overestimated when attenuation correction is not applied (6,7).

In contrast, SPECT/CT imaging may improve the accuracy of the LSF assessment. According to the EANM standard operational procedure on “a unified methodology for ^99m^Tc-MAA pre- and ^90^Y peri-therapy dosimetry in liver radioembolization with ^90^Y microspheres” patients with substantial lung shunt visible in planar imaging are recommended to have an additional SPECT/CT scan performed to ensure a more accurate quantification of the LSF (3).

Regardless of which ^99m^Tc-MAA imaging method (planar or SPECT) is used to predict the distribution of ^90^Y microspheres, there are a number of factors affecting the mismatch between the simulation with MAA and the actual therapy distribution, the most important of which are: differences in shape and size distribution between MAA particles and therapeutic ones, different number of injected particles between the ^99m^Tc-MAA simulation and the therapy session, and uncertainty about the stability of MAA after labelling (3,8,9).

Another option that has been proposed is to inject the patient with the so called *scout dose* consisting of a small batch of microspheres, identical to those used for treatment. The goal is to better simulate the treatment (10). However, the pretreatment activity of β^−^ emitters, used normally to destroy diseased tissue, must be limited. For ^90^Y the estimated safety threshold is about 100 MBq (5,8,10). Of course, such low activity makes imaging even more challenging.

In this study we aimed to assess the accuracy of LSF estimation by means of phantom imaging. To this end we analysed both high activity ^90^Y posttreatment scans and pretreatment scans with the isotope activity of ∼100 MBq, using different nuclear imaging techniques: PET/CT, Bremsstrahlung SPECT/CT as well as planar imaging.

^90^Y imaging using gamma camera (both planar and SPECT) is based on registration of Bremsstrahlung in a wide energy window. Therefore, it poses a major problem when exact attenuation or scatter correction is to be applied (11). Although dedicated Monte Carlo-based reconstruction algorithms have been developed (12) for which a very good quantitative accuracy of the imaging was reported, they are still very time consuming and not widely available. Additionally, it is worth emphasizing that planar imaging has its own inherent limitations, when it comes to quantitative analysis (6,13,14). Regarding SPECT, we tried to optimise the acquisition protocol for ^90^Y imaging by acquiring data with different energy window settings and postprocessing.

For PET, the very low positron emission probability is the main fundamental limit to image quality (11). The small branching ratio related to the internal pair production during ^90^Y decay, directly related to the acquired number of coincidences, may be insufficient to accurately estimate lung shunting, especially at low activities of ^90^Y (8). However, even in high activity ^90^Y posttreatment scans, activity concentration in the lung area may be very low, which makes its quantification challenging due to scarcity of registered counts.

Based on our previous experiences with ^90^Y phantom studies (11,15), we continued the topic of visualization of hot and cold foci in the phantom for different isotope concentrations. The use of an anthropomorphic phantom (instead of Jaszczak or NEMA phantoms) with liver insert containing fillable spheres allowed us to approximate conditions similar to clinical ones. As in our previous study, tumour visibility analysis based on PET/CT and SPECT/CT imaging data was performed both qualitatively and quantitatively using contrast-to-noise ratio as a quantitative parameter (11). Calculated quantitative measure was then compared to the results of qualitative assessments. With the clinical aspect of this current work in mind, we also simulated an extrahepatic lesion in order to investigate whether it can be detected using hybrid nuclear imaging techniques, in both ^90^Y post- and pretreatment scans.

## Materials and Methods

### Phantom

An anthropomorphic torso phantom (model ECT/TOR/P) with a cardiac insert (model ECT/CAR/I) was used to simulate a clinical setting. The phantom included a cylindrical spine insert, a fillable liver insert with two fillable spheres (to simulate both cold - with no activity, and hot spherical intrahepatic tumour), and lung inserts containing styrofoam beads to simulate lung tissue density. The diameter of the hot sphere was 17 mm, and of the cold one - 22 mm. The liver and lungs compartments were filled with ^90^Y chloride to simulate an LSF of 9.8%. The initial activity in the liver was 1451 MBq (including 19.4 MBq in the hot tumour, which meant that the tumour to background ratio was 7.9). In the lungs the activity was 158 MBq. The cardiac insert including a fillable defect of the volume of 2.6 ml was used to simulate an extrahepatic deposition located in between the lungs, above the liver. The defect was filled with an initial ^90^Y activity of 9.8 MBq. The remainder of both the anthropomorphic phantom and cardiac insert were filled with water.

Before phantom experiments with ^90^Y, all fillable compartments of the anthropomorphic phantom as well as the fillable cardiac defect were rinsed with a non-radioactive yttrium chloride solution in 0.5 M of hydrochloric acid to prevent adhesion of ^90^Y to the plastic phantom walls.

### Image acquisition and reconstruction

Nine measurement sessions including PET/CT, SPECT/CT and planar imaging were performed at activities in the whole phantom ranging from 1618 MBq down to 43 MBq. The last imaging session was performed on the 14^th^ day after phantom filling. Total activities in the phantom at the time of PET, SPECT, and planar imaging are listed in Table 1. Because both SPECT and planar images were acquired more than once during each measurement session (as described later), the activity listed in the table is that of the first scan of each of those modalities.

**Table 1.** Phantom activities at the time of PET, SPECT, and planar acquisition.

|  | Days after phantom filling |  |  |  |  |  |  |  |  |
| --- | --- | --- | --- | --- | --- | --- | --- | --- | --- |
|  | 0 | 1 | 3 | 4 | 7 | 10 | 11 | 12 | 14 |
|  | Total <sup>90</sup>Y activity in the phantom [MBq] |  |  |  |  |  |  |  |  |
| PET acquisition | 1618 | 1258 | 746 | 578 | 266 | 121 | 94 | 80 | 43 |
| SPECT acquisition | 1581 | 1275 | 766 | 583 | 272 | 127 | 99 | 78 | 43 |
| Planar acquisition | 1591 | 1285 | 773 | 592 | 271 | 126 | 95 | 78 | 44 |

### PET/CT imaging

PET/CT imaging was performed with a GE Discovery 710 scanner. First, a scout view and a low-dose 64-slice CT scan were performed for attenuation correction of emission data and localisation of the phantom structures. CT scan was acquired with a tube voltage of 140 kV in the helical mode with a current modulation in the range of 40–120 mA. The X-ray tube rotation time was 0.8 s. The helical thickness was 3.75 mm. For the standard type of reconstruction the slice thickness was 1.25 mm. The matrix size was 512×512 (11).

Following CT, three-dimensional PET images were acquired using clinically applied protocol with acquisition time of 30 min per bed position (15.7 cm with 23% bed overlap). Two bed positions were scanned to fit the entire anthropomorphic phantom in the axial field of view (FOV).

Emission data was corrected for geometrical response, detector efficiency, system dead time, random coincidences, scatter and attenuation. Attenuation corrected images were obtained with the use of 3D-OSEM iterative reconstruction method. It was conducted with TOF PET reconstruction algorithm and a resolution recovery algorithm with 4 iterations/32 subsets and a filter cut-off of 3.0 mm. The matrix size was 256×256 (11).

Considering the relatively long acquisition time on PET/CT scanner, a correction for the natural background was applied by performing a series of three PET measurements with the anthropomorphic phantom containing no activity. The PET/CT data was acquired on different days. Each time the settings described above were used.

### SPECT/CT imaging

SPECT/CT images were acquired on a hybrid dual-head GE Infinia VCHWK4 gamma camera with HEGP collimators mounted. A single SPECT acquisition was enough to image the entire phantom in the FOV. For each imaging session Bremsstrahlung SPECT was performed three times with different energy window settings, which are shown in Table 2. One of the energy windows, labelled as W3, was divided into 4 narrower emission windows. This was to provide more accurate attenuation correction based on CT scans. Because different emission energies were acquired in separate sets, a separate CT-based attenuation map was produced for each energy. Then, each set was reconstructed individually and reconstructed images were summed up.

**Table 2.**
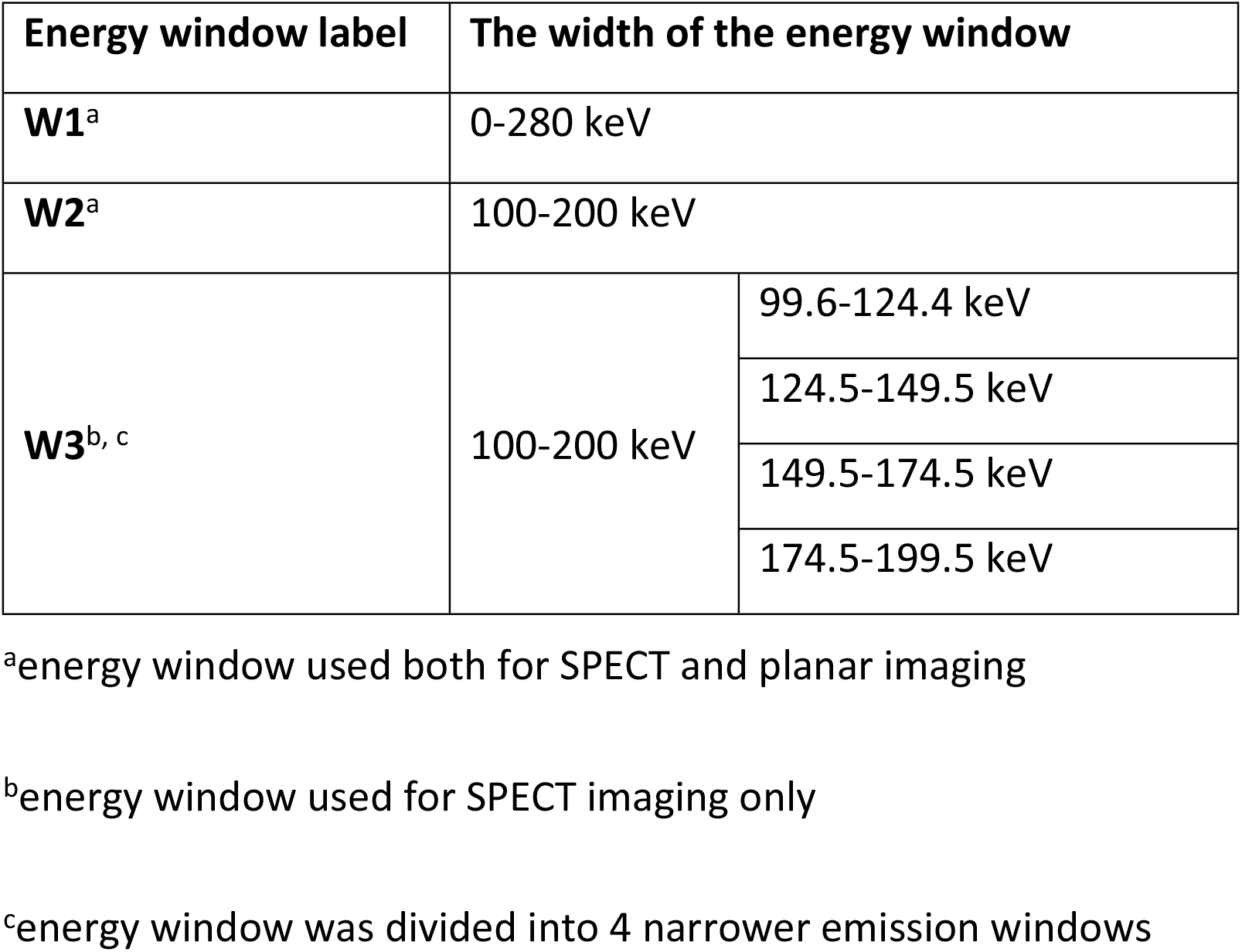
Energy window settings for SPECT and planar acquisition.

| Energy window label | The width of the energy window |  |
| --- | --- | --- |
| **W1<sup>a</sup>** | 0-280 keV |  |
| **W2<sup>a</sup>** | 100-200 keV |  |
| **W3<sup>b, c</sup>** | 100-200 keV | 99.6-124.4 keV |
|  |  | 124.5-149.5 keV |
|  |  | 149.5-174.5 keV |
|  |  | 174.5-199.5 keV |

<sup>a</sup>energy window used both for SPECT and planar imaging
<sup>b</sup>energy window used for SPECT imaging only
<sup>c</sup>energy window was divided into 4 narrower emission windows

For each scan 60 projections were acquired in step and shoot mode, with the angular step of 6°. Total angular range was 360°. Body-contour orbit was used to keep the camera detectors close to the phantom. The acquisition time was set for 30 seconds per projection. The matrix size was 128×128 with a pixel size of 4.42 mm. OSEM reconstruction with 2 iterations and 15 subsets was used. Butterworth filter with a cut-off frequency of 0.5 cycles/cm and a power of 10 was used as a 3D postfilter. No scatter correction or resolution recovery algorithm was applied. Following emission tomography, a CT scan was performed in the axial mode with a tube voltage of 140 kV and a current of 5 mA. The matrix size was 512×512 (11). Similarly to PET/CT, the CT scan was made for attenuation correction and to support organ delineation (8).

### Planar imaging

Planar scintigraphy was performed by means of static scans with acquisition time of 5 min and the matrix size of 256×256. Two detectors were set in H-mode to acquire anterior and posterior images. For each series of measurements the static images were acquired twice using energy windows of W1 and W2. Same as for SPECT imaging, the HEGP collimators were used.

## Data analysis

### Lung Shunt Fraction estimation

Q. Volumetrix MI application on Xeleris 4.1 XFL was used for organ segmentation and quantification for PET reconstructed images.

For PET imaging the liver and lung VOI were manually delineated on one CT scan, referred to as “reference scan”, which in turn was rigidly registered to all other CT scans. All VOIs were transformed accordingly from the reference scan to the other CT scans (5,8). The LSF was calculated directly as the activity in the lungs divided by the activity in the liver and lungs.

Additionally, for PET imaging an extra region without activity within the phantom was delineated to simulate an LSF of 0% (LSF_simulated_). Since its volume was smaller than that of the lungs, the activity in the cold region was re-scaled to avoid underestimation of the LSF_simulated_. The VOI of the cold region was transformed as described above.

To include a correction for the natural background in PET imaging, the mean activity concentration expressed in MBq/ml for the whole phantom volume was first determined, and then the background activity both for liver and lung volume was calculated. The LSF_BKG_corrected_ was then calculated as the activity in the lung VOI divided by the total activity in the liver and lung VOI, whereby the ^90^Y activity in the VOI was reduced by the value of background activity in the corresponding VOI.

SPECT data were reconstructed with the use of Q. Volumetrix MI software on Xeleris 4.1 XFL workstation. However, for organ segmentation and quantification Q.Volumetrix AI available on Xeleris V was used.

To calculate the LSF based on SPECT images there was no need to convert “counts per pixel” to the activity concentration. Since counts are assumed to be proportional to activity, the LSF can be calculated as the counts from the lung VOI divided by the total counts for the liver and lung VOIs.

The methods for organ segmentation as well as VOI transformation between different CT scans were similar to those used for PET. The LSF of 0% was simulated in a similar manner as in PET.

Planar data was processed and analyzed on Xeleris 4.1 XFL workstation using basic applications.

For planar imaging the conjugative view technique was used. A single geometric mean image, composed from the anterior and flipped posterior projection scans, was created each time, and the data analysis was based on geometric mean counts for each ROI (2). Two-dimensional organ delineation was partially supported by a CT scan acquired from hybrid SPECT/CT acquisition. Both the liver and lung ROI were manually delineated based on additional 70.69-mm thick coronal views of a CT scan. The 2D ROIs created in this way were then copied into planar images and position adjusted. For each series of measurements, the phantom was not moved between the planar and SPECT/CT imaging.

We used two approaches to calculate the LSF value. The first was to directly calculate the LSF as defined: the counts from the lung ROI divided by the total counts for the liver and lung ROIs.

The second approach included background correction (3). The method required additional background ROIs to be drawn close to the corresponding organ ROIs. The net number of counts in the liver or lungs N_organ_ was then obtained according to equation:

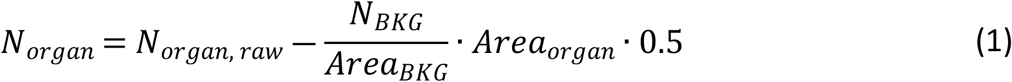

where N_organ, raw_ is the number of counts in the organ ROI, N_BKG_ is the number of counts in the associated background ROI, Area (Area_organ_, Area_BKG_) is the size of the ROI, and 0.5 is a correction factor for background, applicable for large organs (3).

### Tumour visibility analysis

In order to assess the feasibility of imaging tumours in the liver both qualitative and quantitative analysis of the data was performed.

Three observers with at least 2 years of experience in SPECT and PET analysed the data qualitatively. For SPECT they reviewed the images acquired for all the imaging sessions and all energy windows settings resulting in 27 datasets, and for PET imaging all 9 timepoints were assessed. The observers were asked to mark the three foci (the hot and cold tumours in the liver as well as the extrahepatic deposition) as either visible or not in all of the datasets.

Quantitative analysis was conducted for the hot and cold tumours in the liver. Lesion detection is heavily impacted by both lesion to background contrast and noise in the image, so contrast-to-noise ratio (CNR) was chosen as a quantifiable parameter of tumour discernibility (11). The Rose criterion states that an object can be detected in the image if its CNR is over a certain threshold, which is usually between 3 and 5 (16).

As in our previously published work, we used an in-house software to define the lesions based on CT images in their widest cross-section. Moreover, we defined background regions close to the analysed foci. We gathered the basic statistics for all chosen regions (including mean values and standard deviations) by transferring the contours onto the nuclear medicine images. CNR calculations were conducted as previously described for NEMA and Jaszczak phantoms (11).

Due to noise in PET/CT data all of those images were analysed after applying Wiener filter (PSF = 5, noise to signal ratio = 0.11).

## Results

### Lung Shunt Fraction estimation

Fig 1 shows the 3D MIP (Maximum Intensity Projection) of the PET and SPECT images of the anthropomorphic phantom for scans performed at high and low activity levels of ^90^Y, whereas Fig 2 shows the geometric mean images enabling the visual assessment of lung shunt in planar imaging. Both SPECT and planar data presented in the figures were acquired using energy window of 100-200 keV (W2). PET images acquired at high activity level clearly showed the presence of the lung shunt of a true LSF of around 10%. However, for low activities (below approximately 200 MBq), the lungs and liver were no longer identifiable in PET reconstructions. In SPECT images, the lung shunt was not visible, even at a therapeutic activity level of about 1.5 GBq. For planar imaging, one could identified visually the lung shunt only in images obtained with high ^90^Y activities.

**Fig 1.**
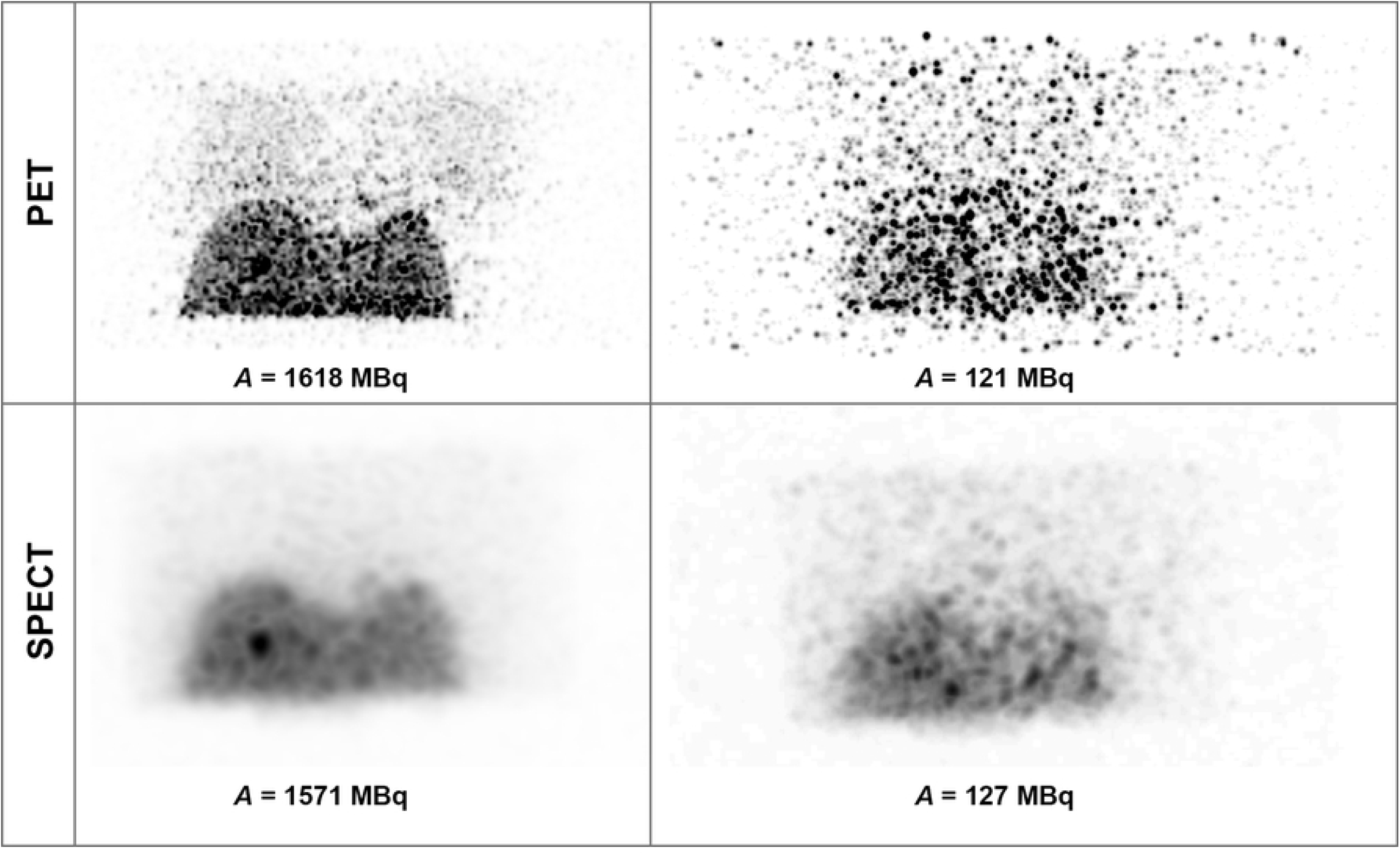
3D MIP PET (top row) and SPECT (bottom row) images of the anthropomorphic phantom for scans performed at high (left) and low (right) activity levels of ^90^Y.

**Fig 2.**
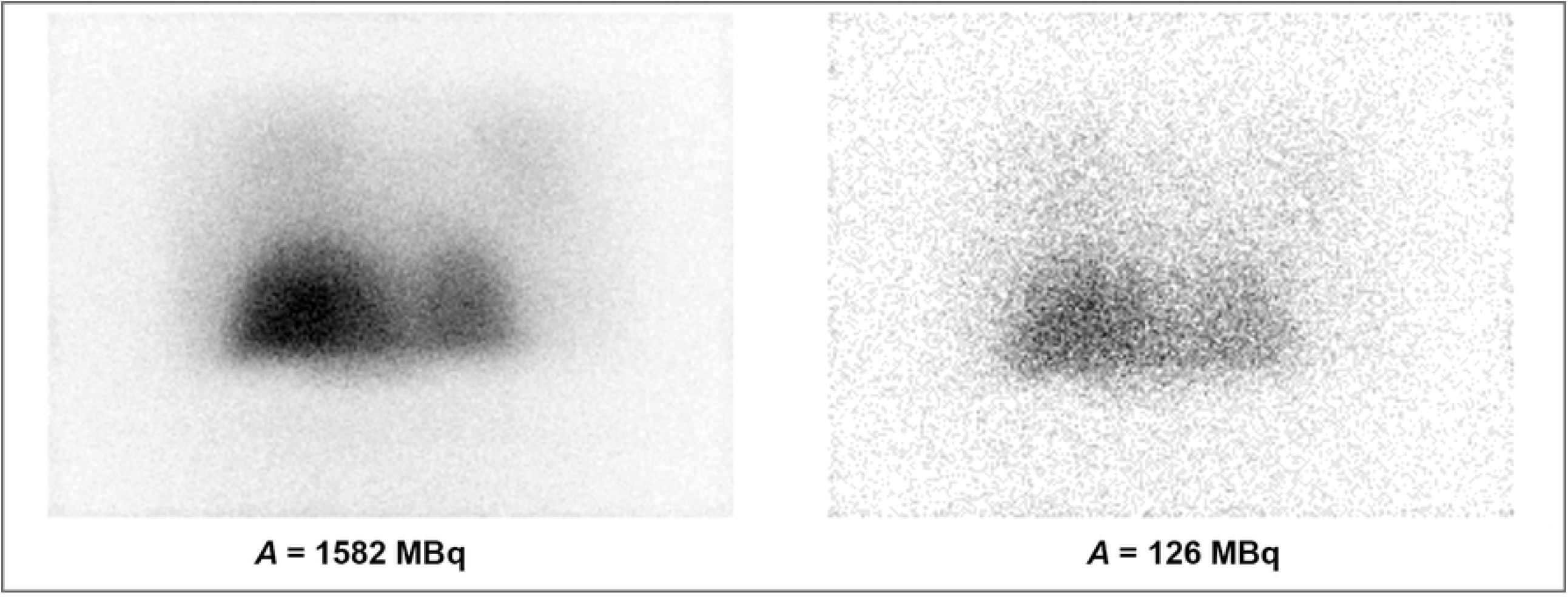
Geometric mean images composed from the anterior and posterior planar images for acquisitions with a total phantom activity of 1582 and 126 MBq.

Figs 3 and 4 show the LSF estimated from PET, SPECT and planar imaging as a function of total phantom activity.

**Fig 3.**
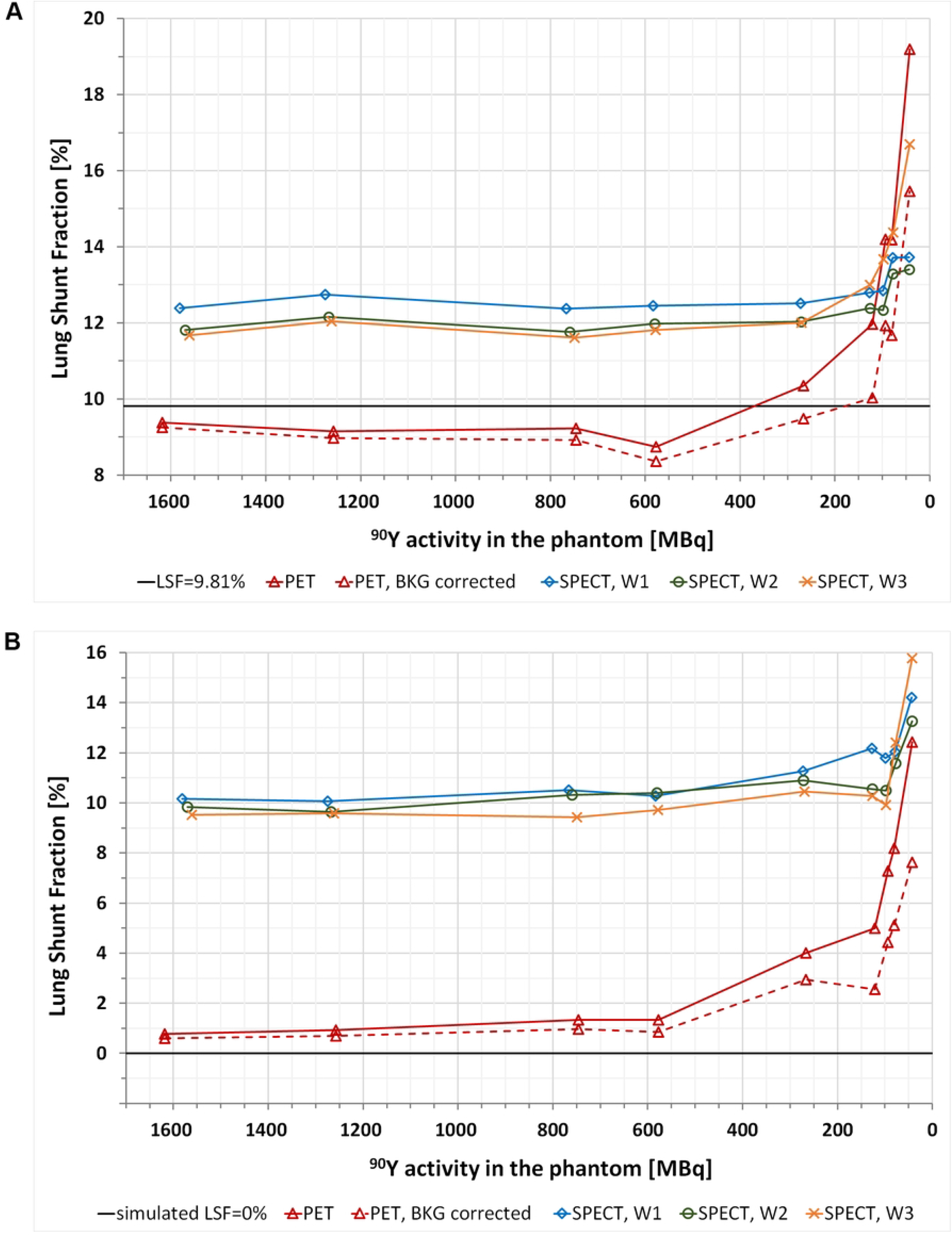
LSF estimated from PET and SPECT imaging for a true LSF of 9.81% (A) and LSF_simulated_, i.e. 0% (B) as a function of total ^90^Y activity in the anthropomorphic phantom. The results for PET data are presented for calculations both with and without the natural background correction. For SPECT modality the results are shown for acquisitions with different energy window settings: W1, W2, and W3.

**Fig 4.**
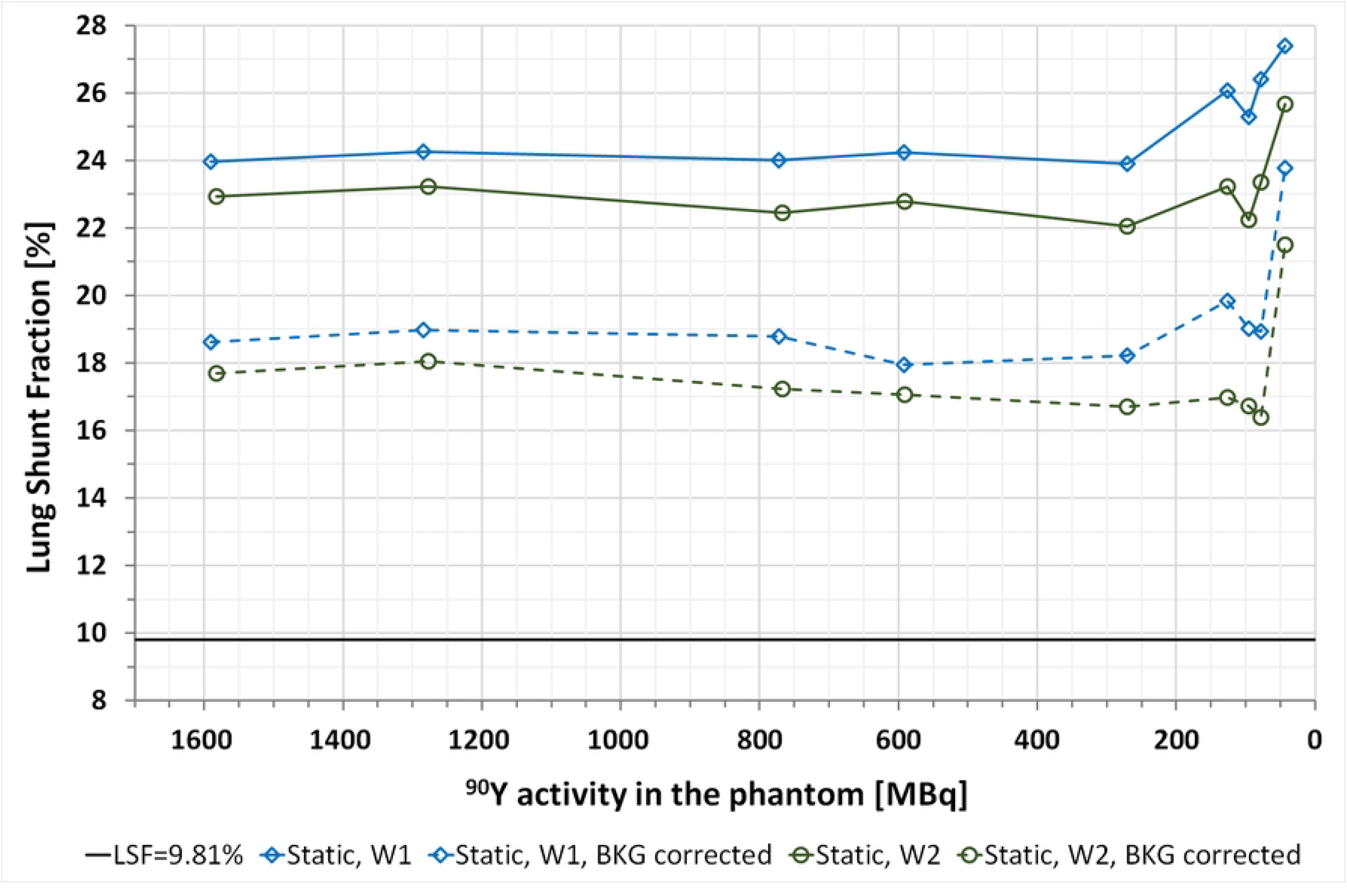
LSF estimated from planar images acquired using energy windows of W1 and W2 for a true LSF of 9.81% as a function of total ^90^Y activity in the anthropomorphic phantom. The results are presented for calculations both with and without background correction.

### PET/CT

For PET imaging the LSF was accurately estimated at activities in the phantom ranging from 1618 MBq down to about 200 MBq (absolute difference less than 1.5pp). However, the LSF estimation was most stable (although slightly underestimated) over the range of high activities down to about 400 MBq. For lower activities, the increase in the LSF value was observed leading to overestimation of the LSF by 2.2pp at 120 MBq, although the background correction allowed to reduce it to 0.22pp. Below 100 MBq, the LSF was largely overestimated (absolute differences of 4.4pp and 2.1pp for PET without and with background correction at 94 MBq). These results are presented in Fig 3A.

The LSF_simulated_, based on the VOI with no activity, was stable over the range of high activities down to approximately 500 MBq, and its value was then less than 1% (when the background correction was applied). However, along with the decrease in phantom activity, the calculated, background corrected LSF_simulated_ increased to about 4.0% and 7.6% at 100 MBq and 45 MBq, respectively. In the range of low activities the true LSF value of 0% was even more overestimated when assessed using PET without background correction (up to 12.4% at 43 MBq). These results are presented in Fig 3B.

### Bremsstrahlung SPECT/CT

Bremsstrahlung SPECT overestimated the LSF regardless of the energy window setting in the whole range of activities. For activities over 200 MBq the largest overestimation of the LSF was present for SPECT when the widest energy window was used (W1). Mean LSF value from the first seven measurements (i.e. for the range of activity where the LSF was relatively constant) was 12.6%. For data acquired with W2 energy window it was 12.1% (with maximum absolute difference of 2.6pp). The W3 energy window yielded results very similar to these of W2 in the range of high and medium activities, however, for lower activities, it resulted in the largest overestimation of the LSF (up to nearly 17% at 43 MBq). These results are presented in Fig 3A.

The estimated LSF for a true LSF of 0% was greatly overestimated. Mean LSF_simulated_ values from the first seven measurements were: 10.9%, 10.3%, and 9.9% for energy windows of W1, W2, and W3, respectively (Fig 3B)

### Planar imaging

Planar images greatly overestimated the LSF (up to 17.6pp for W1 and 15.9pp for W2). The background correction reduced the LSF overestimation by approximately 5.7pp and 5.5pp for the W1 and W2 energy window, respectively (Fig 4).

## Foci visibility

The reconstructed images (both PET and SPECT) were used for qualitative and quantitative analysis. The observers used both coronal and axial views to determine the visibility of the foci. They could review single modality as well as fused images. On the other hand, the CNR calculations were performed on the axial views of the phantom (Fig 5).

**Fig 5.**
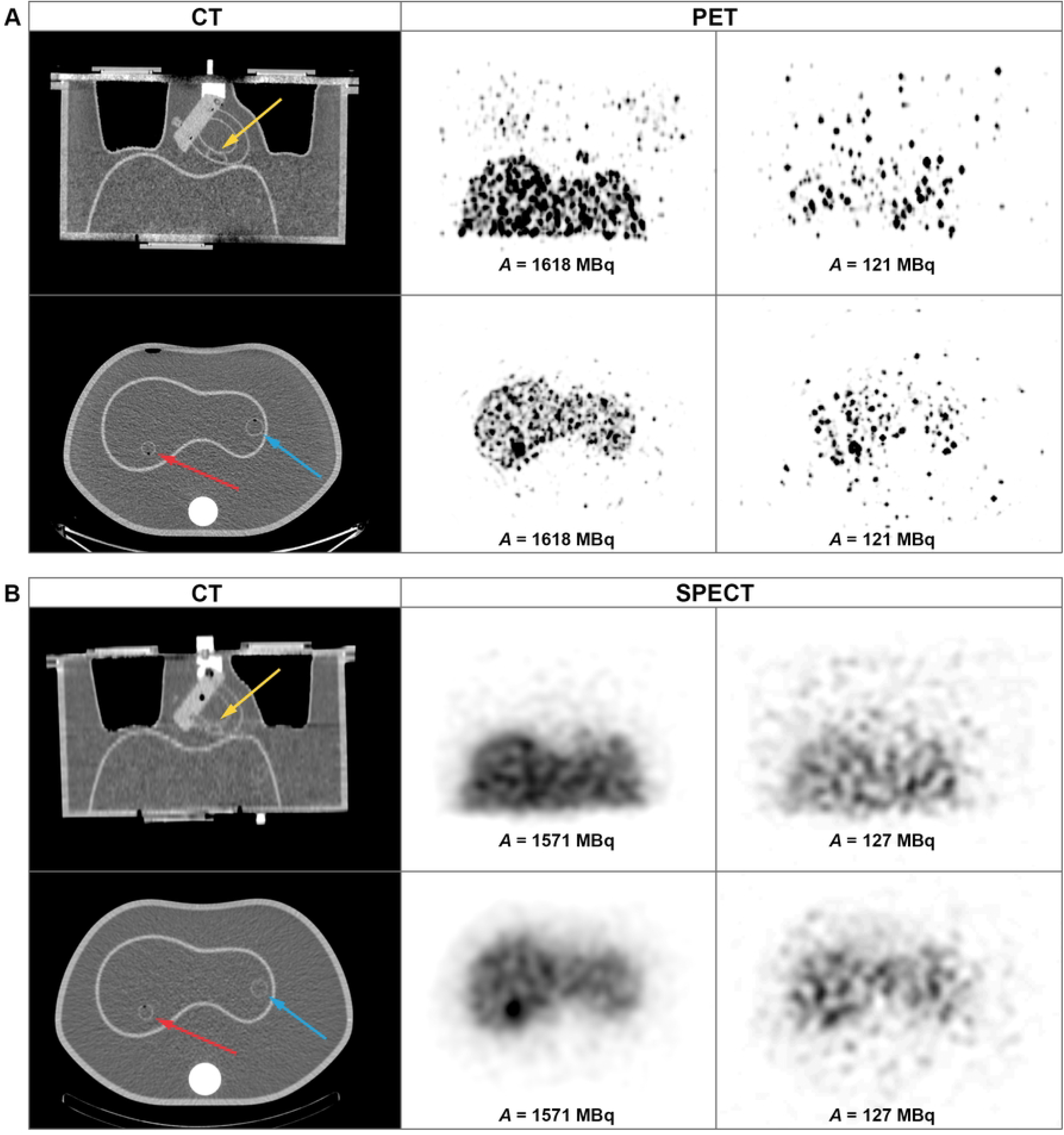
Coronal (top row) and axial (bottom row) views of the PET (A) and SPECT (B) images of the anthropomorphic phantom for scans performed at high and low activity levels of ^90^Y (middle and right column). Left column presents corresponding CT slices. Yellow arrows indicate the location of the extrahepatic deposition while the red and blue ones - the locations of the hot and cold hepatic tumours, respectively.

### SPECT/CT

In the qualitative assessment of SPECT/CT images the hot lesion was visible down to the activity in the whole phantom of 266 MBq (liver - 239 MBq, hot tumour - 3.2 MBq, with tumour to background ratio of 7.9). However the cold sphere proved not to be visible in any of the acquired images.

The W1 and W2 energy windows acquisitions yielded very good agreement between qualitative and quantitative assessment of foci visibility: CNRs calculated for visible lesions were above 3, which is one of the border values suggested by Rose criterion. For results obtained from the W3 energy window images the agreement was not satisfactory, as the CNRs of the hot lesion acquired from the first two timepoints were below 3. For the cold lesion, which could not be distinguished in any of the images, the CNR was well below 3. These results are presented in Figs 6 and 7.

**Fig 6.**
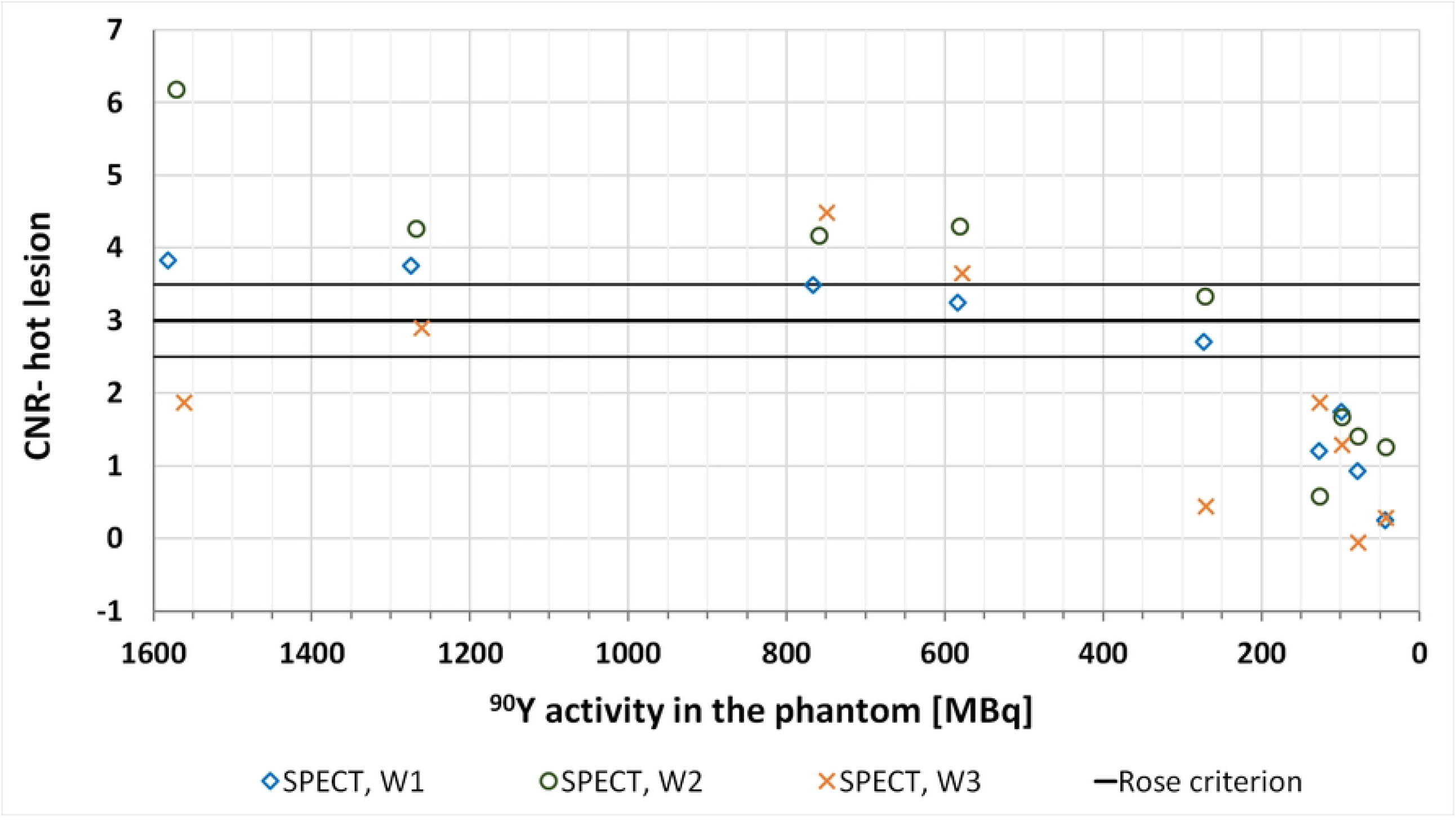
CNR values calculated for the hot lesion in the liver in SPECT/CT imaging for all of the analysed energy window settings. The solid lines represent the border values depending on the Rose criterion (middle line at 3 and supporting ones at 2.5 and 3.5).

**Fig 7.**
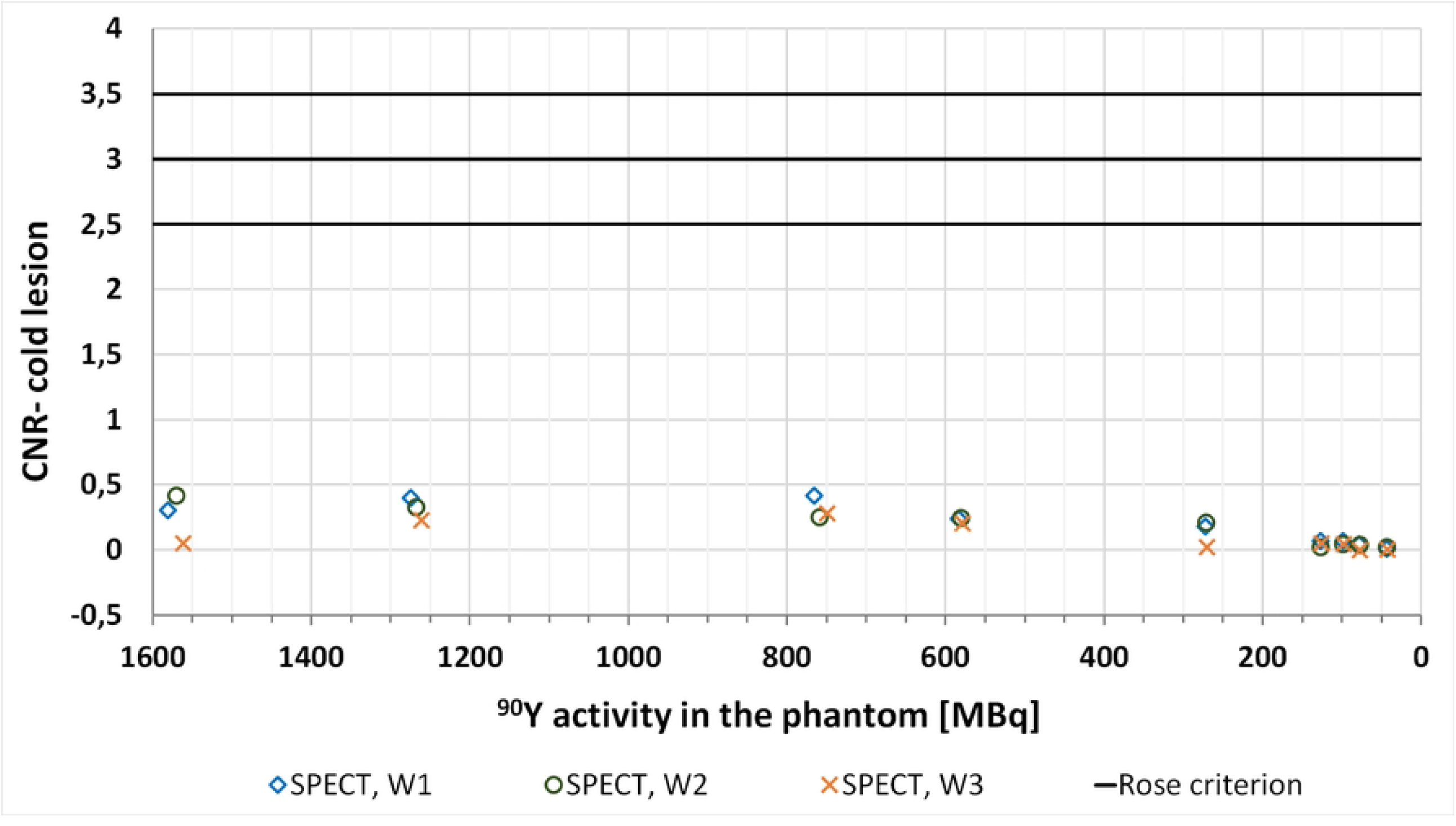
CNR values calculated for the cold lesion in the liver in SPECT/CT for all of the analysed energy window settings. The solid lines represent the border values depending on the Rose criterion (middle line at 3 and supporting ones at 2.5 and 3.5).

We have also qualitatively analysed the visibility of the extrahepatic concentration (Fig 5). It remained visible up to the 3^rd^ day after phantom filling, when activity in the whole phantom was 747 MBq (in the extrahepatic deposition the activity was then 4.5 MBq). However, it needs to be noted that the last dataset with discernible activity concentration outside of the liver required a careful review of all available cross sections.

### PET/CT

In the qualitative assessment of the PET/CT data the cold focus was visible in the first acquired dataset, whereas the hot lesion was discernible up to the ninth day after the filling of the phantom (activity in the tumour at this point was 1.46 MBq). These results were in agreement with the calculated CNR values, which for the visible tumours were mostly over the border value of 3 as suggested by the Rose criterion (Fig 8).

**Fig 8.**
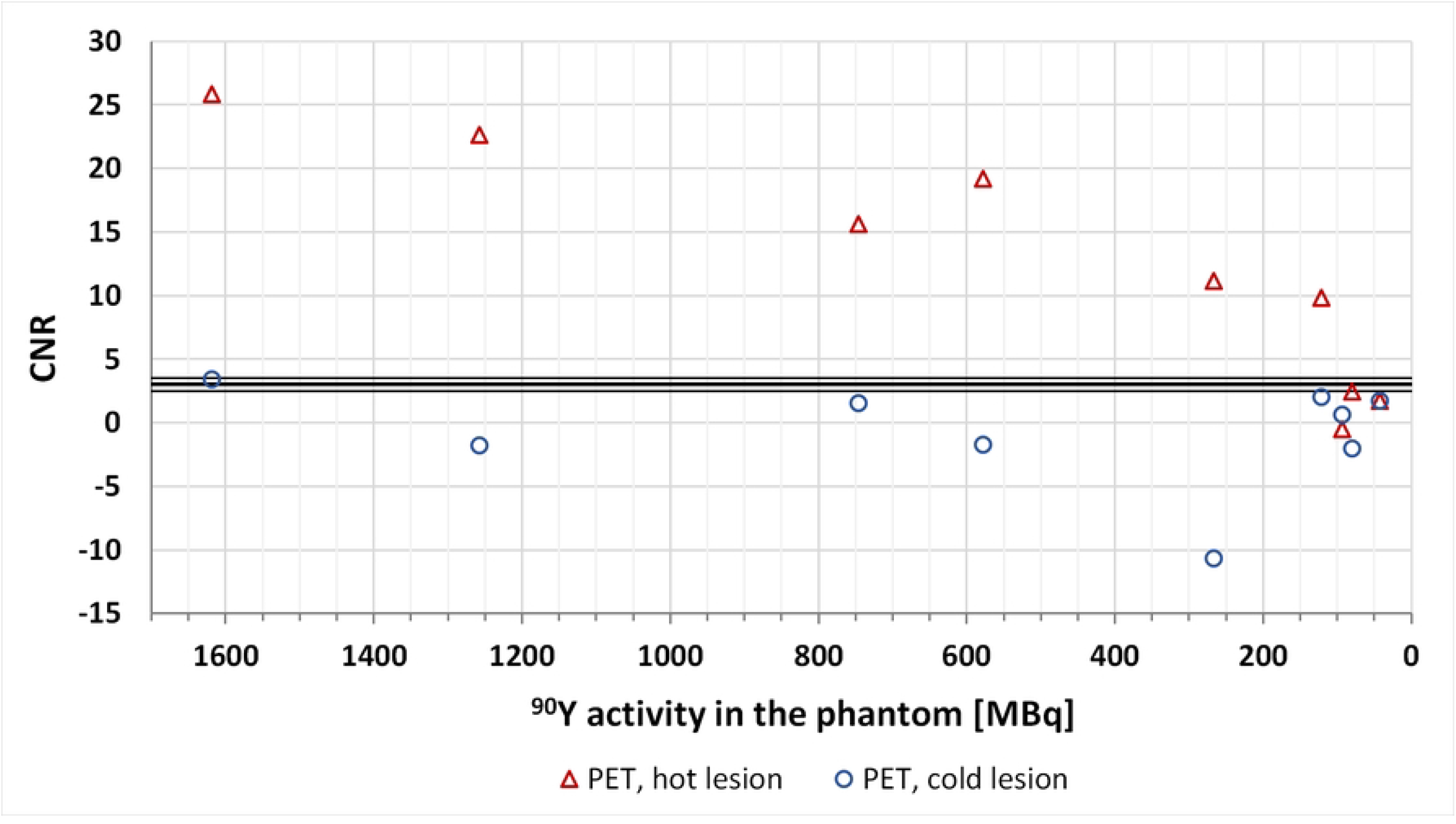
CNR values calculated for the cold and hot lesions in the liver in PET/CT imaging. The solid lines represent the border values depending on the Rose criterion (middle line at 3 and supporting ones at 2.5 and 3.5).

As with SPECT, the observers were able to differentiate the extrahepatic lesion up to the 3^rd^ day after phantom filling.

## Discussion

This study is a continuation of our previous work (11), in which we analysed the feasibility of SPECT/CT and PET/CT ^90^Y imaging. In this study, we aimed at approximating clinical applications not only by using an anthropomorphic phantom (as opposed to NEMA and Jaszczak phantoms), but also by analysing a pretreatment parameter used to qualify patients for therapy with ^90^Y microspheres. In this work we have assessed the accuracy of LSF estimation, the feasibility of imaging hot and cold hepatic tumours as well as extrahepatic lesions using different imaging modalities.

For PET/CT images we are able to visually distinguish the organs of interest (liver and lungs) for activities in the phantom over 200 MBq. For lower activities the PET reconstructions are very noisy leading to poor discernibility of the liver, while the lungs cannot be identified at all (Fig 1). On the other hand, in SPECT images (both in MIP and cross-sectional planes) the lung shunt is not visible, even for high activities (Figs 1 and 5). The liver is easily identifiable on almost all scans. However, the spill-over effect of activity outside of the liver cannot be ignored, as it adversely affects the possibility of accurate estimation of activity in the background and neighbouring organs. This effect is also connected to lower spatial resolution than in PET technique. Similar spill-over effect could be observed in the planar images. In those images the liver was as distinguishable as in SPECT, however the lungs were identifiable in images obtained with high activities. It has to be noted, that the visibility of lungs was worse than in PET reconstructions. Both SPECT and planar imaging was strongly influenced by scattered radiation, due to registration of photons in wide energy windows. Neither of those techniques included corrections for this effect, which adversely affects the quality of those images. These qualitative findings are in good agreement with results of study by Kunnen et al. (8), even though they used a higher LSF value of 15%.

We chose the lung shunt value of 10% because for resin microspheres (SIR-Sphere) it is the experts’ recommended lung shunt threshold beyond which a reduction in isotope activity is strongly advised (1,17,18). Therefore, the LSF of about 10% needs to be detectable and accurately estimated.

The PET data yielded the most accurate LSF estimation as presented in Fig 3A. In our study the absolute differences of calculated and true LSF values for total activities in the phantom over 200 MBq were less than 1.5pp. In comparison, Kunnen et al. report differences of less than 2pp for similar activity range. Similarly to the aforementioned study, we also have observed that background correction had the most effect on LSF calculations for lower activities (less than 100 MBq) (8). The same author published another study, in which they analysed lower LSF of 5.3%. They show poor estimation of LSF for activities below 1 GBq, for calculations based on PET images (5). This might suggest that LSF below 10% might be difficult to properly assess in PET, and indicates a need for further study on this subject.

For SPECT data the LSF was overestimated and was between 12% to 13% for activities over 200 MBq. Even though this seems like a rather good estimation, it cannot be overlooked, that the same method yielded LSF of about 10% for cold VOIs used to simulate LSF of 0% (Fig 3B). This quantitative result correlates well with the qualitative, visual assessment of the SPECT/CT images, in which the lungs filled with activity did not differ from the cold background. It is worth noting, that LSF obtained by Kunnen et al. using similar clinical acquisition protocol were overestimating the real values by a much higher margin (13pp) at high activities.

Unlike SPECT, the calculated LSF for a true LSF of 0% for PET was only slightly overestimated (Fig 3B). This suggests that the obtained results are much more reliable than the values calculated from SPECT data.

In planar imaging we observed a gross overestimation of calculated LSF (Fig 4). The background correction performed according to EANM recommendations (3), allowed for its partial reduction. Nevertheless, the obtained results were definitively worse than those from PET or SPECT. In part, it can be explained by the lack of scatter and attenuation correction, as well as overlapping of different structures.

As a continuation of our previous research we have analysed the visibility of hot and cold foci in the liver. We have found that PET acquisitions provided better images for distinguishing the tumours from the liver. The hot lesion was visible even for low activities, while the cold lesion was reported to be visible both qualitatively and quantitatively only for the highest activity of ^90^Y (Fig 5A).

Previously we have demonstrated that the smallest cold foci visible in our SPECT acquisitions of the Jaszczak phantom had a diameter of 25.4 mm (11). Since we wanted to analyse tumours’ visibility at higher concentrations of ^90^Y activity in surrounding area closer to clinical settings, we have chosen to use a slightly smaller sphere (diameter of 22 mm). The cold tumour was not visible in any of the acquired SPECT scans (Fig 5B). This might be explained by poor spatial resolution and spill-over activity from the liver. The acquired CNR values are consistent with the lesion’s discernibility and our previous findings.

We have found good correlation between CNR and hot tumour visibility for energy window W1 and W2, with higher values calculated for the latter. For the W3 energy window there were significant discrepancies. The first two acquisitions, which should have provided the highest CNR values, proved to be below the Rose criterion border value of 3. It might have been influenced by the fact that the images analysed for the W3 energy window session were summed reconstructions, as opposed to W1 and W2 sessions. The choice of energy window seems to have rather minor effect on both the detectability of lesions and the accuracy of LSF estimation.

For SPECT imaging based on registration of Bremsstrahlung reconstructions commonly used in clinical practice inherently suffer from insufficient attenuation correction and lack of scatter correction. These shortcomings might be overcome by implementation of Monte Carlo based reconstruction algorithms (19,20). However, these methods are time and labour consuming and are not easily accessible for many smaller nuclear medicine centres.

Our study suggests that PET/CT provides better images for high activity posttreatment imaging of patients undergoing therapy with ^90^Y microspheres than Bremsstrahlung based SPECT/CT. Lowering the activity to levels acceptable in pretreatment scans (about 100 MBq), poses a great challenge in imaging. Its main goal is to reliably assess the LSF. Our work demonstrates that PET/CT yields the best results among the tested methods. However, it seems that it may have some inherent limitations in imaging (low counts in the image), which explains the difficulties with image interpretation and analysis at the lowest activities. SPECT/CT could be considered for this purpose, if suitable corrections can be applied. As for planar imaging it did not prove to provide enough information for it to be clinically useful for pretreatment ^90^Y scans.

## Data Availability

ll relevant data are within the manuscript and its Supporting Information files.

## Acknowledgments

The authors thank Anna Szarowicz from GE Healthcare for enabling the use of Xeleris V’s new application “Q.Volumetrix AI” in this study for SPECT/CT segmentation and quantitation.

## Supporting information

**S1 File. PET, SPECT and planar data for LSF calculations**. (XLSX)

**S2 File. CNR for SPECT and PET**. (XLSX)

